# Changing pattern in presentation of infective conjunctivitis during COVID-19 pandemic in a tertiary eye hospital in Nepal

**DOI:** 10.1101/2024.10.01.24314750

**Authors:** Thinley, Reeta Gurung, Hom Bahadur Gurung, Manish Poudel, Pradeep Banjara

**Author notes:** **Corresponding Author:** Thinley, General Ophthalmologist, Department of Ophthalmology, Eastern Regional Referral Hospital Mongar, Bhutan., Postal code: 43001. Department of Ophthalmology, Eastern Regional Referral Hospital, Mongar 43001, Bhutan (Thinley).

## Abstract

**Purpose:** To evaluate changes in the pattern of presentation of infective conjunctivitis during COVID-19 pandemic at a Tertiary Eye Hospital in Nepal.

**Methods:** This was a single-center retrospective cross-sectional study that analyzed cases of infective conjunctivitis at Tilganga Institute of Ophthalmology, Nepal from January 2018 to December 2022 using data from electronic medical records. The data from 2020 and 2021 were considered as the COVID era. Cases of corneal foreign body during the same period were used as a control.

**Results:** The number of cases of infective conjunctivitis decreased by 64.86% (95% CI, -64.82% to -64.94%; p<0.001) during the COVID-19 pandemic. Sharp decrease was noted after the implementation of lockdowns and preventive measures. However, in post-pandemic era, the number of cases increased by 24.6 % (95% CI, +24.57% to +24.66%; P<0.001). In contrast, the cases of corneal foreign body increased by 41.63 % (95% CI, +41.59% to +41.66%; p <0.001).

**Conclusion:** The number of cases of infective conjunctivitis at our hospital decreased significantly during the COVID-19 pandemic. This reduction in number and change in pattern of presentation might be attributed to the widespread implementation of preventive public health measures during the pandemic period.

## Introduction

Infective conjunctivitis is a common eye condition characterized by inflammation of the conjunctiva, typically caused by viral or bacterial infections (1). Acute conjunctivitis, commonly called as pink eye, is highly contagious and very common condition. It is estimated that conjunctivitis accounts for 1% to 2% of primary care visits in the USA (2) and accounts for almost one-third of all eye related visits in emergency departments (3).

Acute viral conjunctivitis is transmitted through close personal contact, fomites, droplets in the air, and poor hand hygiene(4) (5). Despite infection control programs and measures, acute viral conjunctivitis continues to be a source of public health concern and a substantial economic burden (6) (7).

During the COVID-19 pandemic, our tertiary eye hospital in Nepal observed a noticeable reduction in the number of cases presenting with infective conjunctivitis to both emergency and outpatient departments. This reduction in cases was hypothesized to be due to the robust implementation of preventive public health measures such as frequent hand hygiene, wearing face masks, and maintaining social distancing.

This hypothesis is supported by studies that have shown a decrease in the overall incidence of respiratory infections, including conjunctivitis, during the COVID-19 pandemic when these preventive measures were widely practiced (8).These measures have been shown to be effective in reducing the transmission of respiratory viruses, including COVID-19 and adenovirus – a common cause of viral conjunctivitis (9).

Furthermore, there have been reports of conjunctivitis being associated with COVID-19 infection. However, the literature on this association is limited and conflicting. Although it has been reported that viral conjunctivitis can be associated with COVID-19, the causal relationship between ocular manifestations and SARS-COV-2 infection is yet to be elucidated (10) (11) (12).

This study aimed to evaluate the changes in the pattern of presentation of infective conjunctivitis at Tilganga Institute of Ophthalmology (TIO) over the span of five years, distributed into pre-COVID, COVID and post-COVID eras.

## Materials and Methods

This was a retrospective analysis of the data on infective conjunctivitis spanning five years, from January 1st, 2018 to December 31st, 2022. The study searched for data on cases of conjunctivitis presenting to the emergency department (ER) and outpatient departments (OPD) of Tilganga Institute of Ophthalmology, using the Institute’s electronic medical records (EMR). The data was accessed between January 23, 2023 to April 30, 2023. The data thus obtained were cleaned and sorted using Microsoft Excel 2016. The data from the years 2018 and 2019 were considered as that of pre-COVID era; 2020 and 2021 as the COVID era, and that of 2022 as the post-COVID era. For comparison purposes, the data from 2018, 2021, and 2022 were used as that of pre-COVID, COVID and post-COVID eras, respectively.

The study included the cases of infective conjunctivitis, based on the presumptive diagnosis and treatments provided by the treating ophthalmologist or resident doctors. Cases of allergic conjunctivitis and other types of conjunctivitis were excluded.

The cases of corneal foreign body that presented to our hospital during the study period were used as a control.

The data was then transferred to and analyzed using SPSS version 20. T-test was used to compare the data of the COVID-era with that of pre and post-COVID eras. All statistically significant analyses were calculated, and the results are presented in the form of percentage, mean, graphs, and charts. P-value of less than 0.05 was considered statistically significant.

The study was approved by the Institutional Review Board of Tilganga Institute of Ophthalmology (Ref: 20/2022 dated 21^st^ Oct. 2022). Informed consent was not obtained for this study since the data was analyzed anonymously. The study has adhered with the Declarations of Helsinki.

## Results

### Patient demography

Among the total of 16,853 cases of infective conjunctivitis during the study period, 9,476(56.2%) were male and 7,377(44.8%) were female. The mean (SD) age of the patients was 32.9(19.1) years.

In the pre-COVID era, the number of infective conjunctivitis cases visiting our hospital (out of total patient attendance) was 5,235(1.8%) and 5,871(1.9%) in 2018 and 2019, respectively. In the COVID era, the figures deceased to 1,878(1.3%) and 1,396(0.6%) cases in 2020 and 2021 respectively. However, in the post-COVID era (2022), the number of cases increased to 2,313(0.8%).

**Figure 1:**
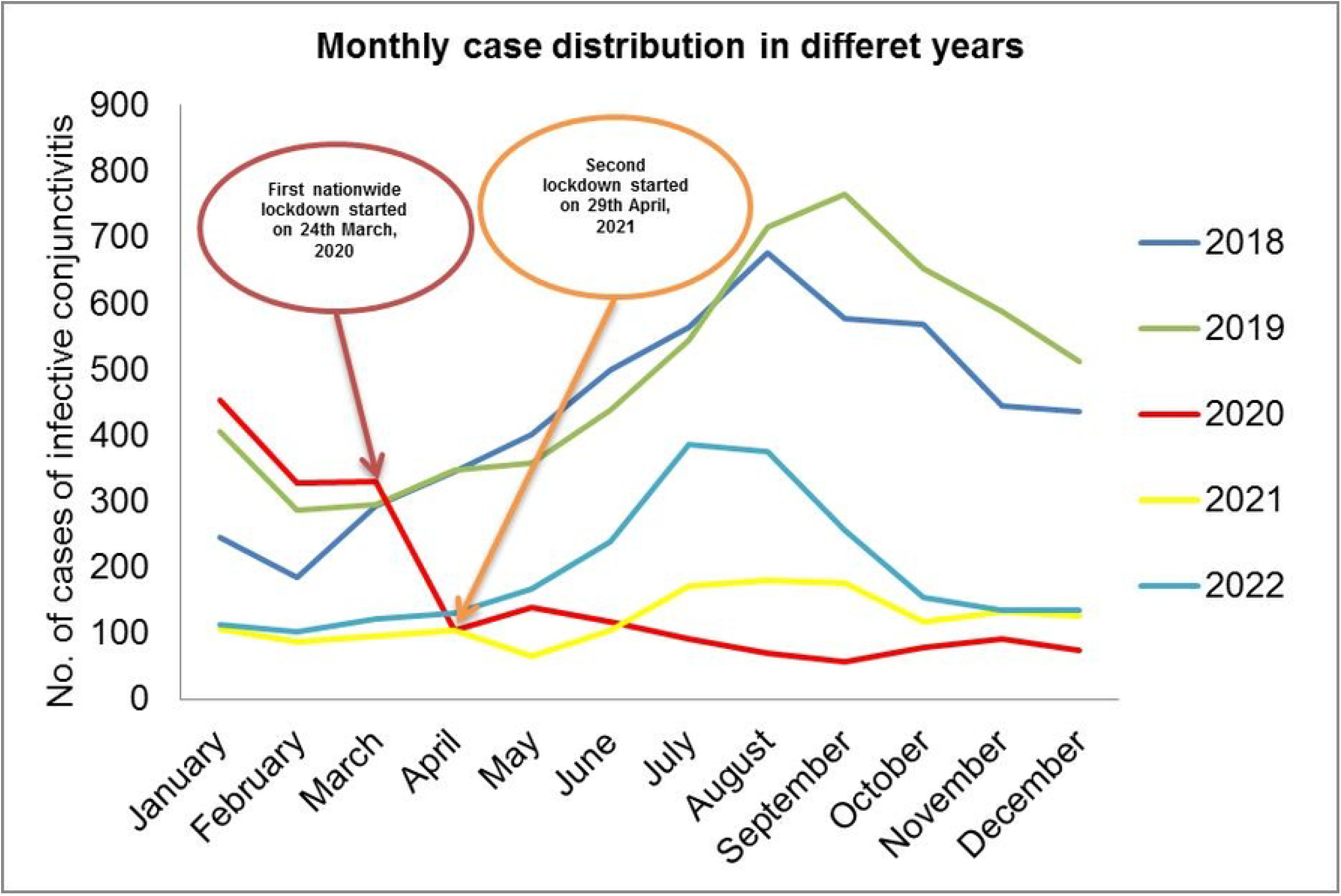
Monthly distribution of infective conjunctivitis cases in different years. The number of cases dropped sharply from April 2020 and remained low throughout the year and a similar pattern continued into the subsequent year (2021).The sharp decline in cases coincided with the implementation of lockdowns. However, in 2022 (post-COVID era), the number of cases began to rise again and followed a monthly distribution pattern similar to that of pre-COVID era.

The mean(SD) monthly cases of infective conjunctivitis presenting to our hospital were 436(142.49) and 492(156.31) in 2018 and 2019(Pre-COVID era), respectively. This number dropped significantly to a mean(SD) of 161(125.94) and 122(34.95) in 2020 and 2021(COVID era), respectively. A consistent trend of increase in the number of cases of infective conjunctivitis was observed from June through October during 2018 and 2019. In contrast, the number of cases decreased sharply from April 2020 and remained low throughout the year, with a monthly mean(SD) of 91.40(23.9), and a similar pattern continued into the subsequent year (2021). The sharp decline in the number of cases coincided with the implementation of lockdowns. However, in 2022(post-COVID era), the number of cases began to rise again and followed a monthly distribution pattern similar to that of pre-COVID era. Nevertheless, the number of cases remained well below the pre-COVID average (**Figure 1**).

The number of cases of infective conjunctivitis presenting to our hospital decreased by 64.86% (95% CI, -64.82% to -64.94%; p< 0.001) during the COVID era. However, in the post-COVID era, the cases increased by 24.62% (95% CI, +24.57% to +24.66%; p<0.001). In contrast, the cases of corneal foreign body showed an increase of 41.63% (95% CI, +41.59% to +41.66%; p<0.001) during the pandemic (**Table 1**).

**Table 1.** Comparison between cases of infective conjunctivitis and corneal foreign body.

| COVID eras | Year | Total no. of Patients | No. of cases of Infective conjunctivitis † | No. of cases of corneal foreign body* | Percentage changes | P-value |
| --- | --- | --- | --- | --- | --- | --- |
| Pre-COVID | 2018 | 282729 | 5233 (1.85) | 1098 (0.39) |  |  |
| COVID | 2021 | 225813 | 1468 (0.65) | 1242 (0.55) | 64.8% ↓ †<br>41.63% ↑ * | <0.001<br><0.001 |
| Post-COVID | 2022 | 286690 | 2313 (0.81) | 1364 (0.48) | 24.6 % ↑ †<br>13.50% ↓ * | <0.001<br><0.001 |

Note: † Infective conjunctivitis
\* Corneal foreign body

**Figure 2:**
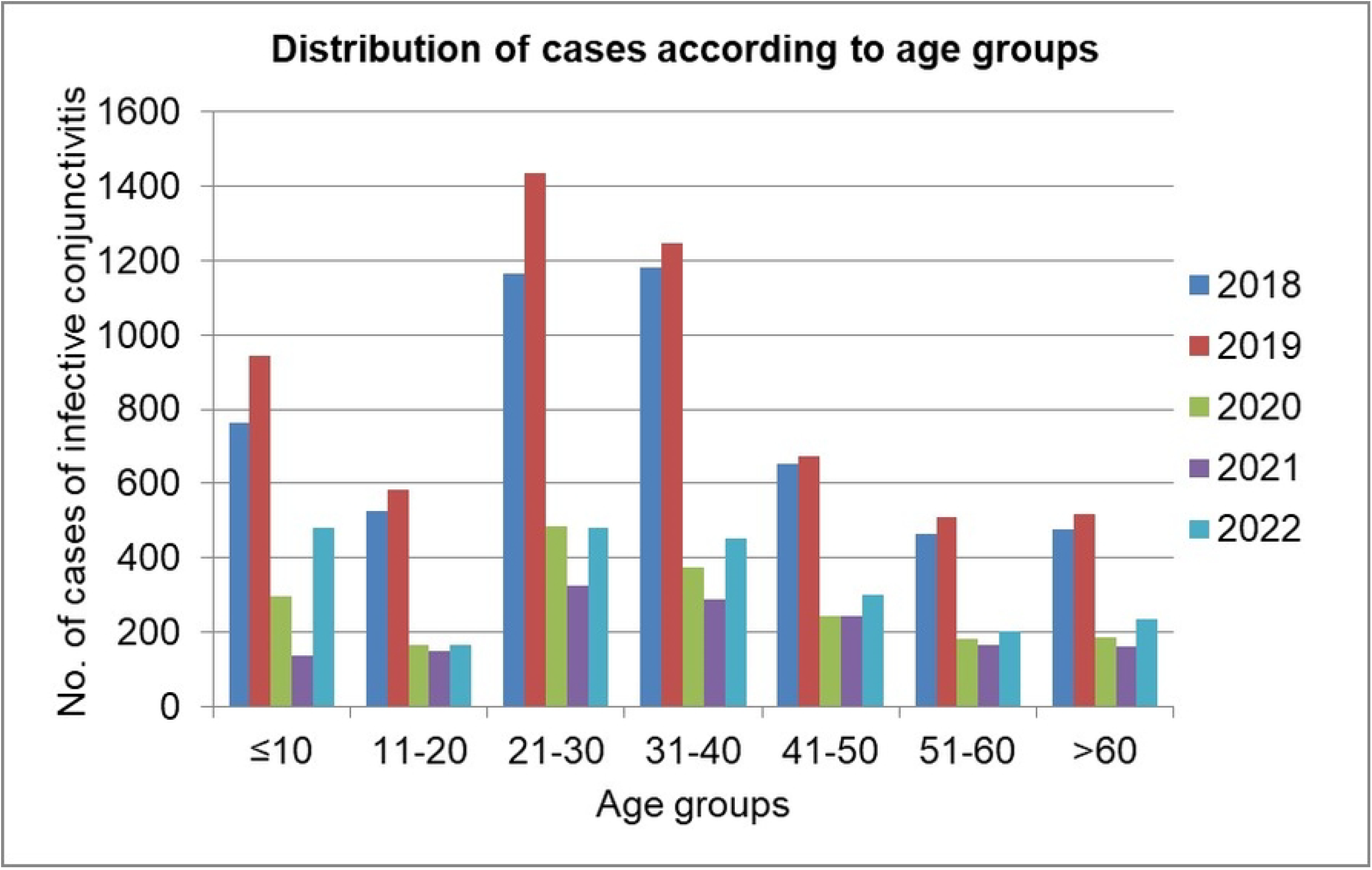
Distribution of infective conjunctivitis cases in different age groups. The most affected age groups are 21-30 and 31-40 years of age. The result also showed that the number of cases of infective conjunctivitis presenting to our center were significantly higher in the pre-COVID era irrespective the age groups.

It was observed that the most affected age groups were 21-30 and 31-40 years of age. The result also showed that the number of cases of infective conjunctivitis presenting to our center were significantly higher in the pre-COVID era irrespective of the age groups (**Fig. 2**).

## Discussion

After the onset of COVID-19 pandemic in December 2019, a noticeable decrease in the number of cases of infective conjunctivitis visiting the emergency and outpatient departments of our hospital was observed. The emergency department of TIO, which usually used to see many cases of conjunctivitis, experienced a perceptible drop during the pandemic period.

The results of our study have shown that the number of cases of infective conjunctivitis presenting to our hospital had decreased significantly during the pandemic. The number of cases of infective conjunctivitis fell by 64.86% during the pandemic era. The decrease in the number of cases correlates well with the time of enforcement of lockdowns. This result may suggest that the public health measures implemented during those time to curb the spread of COVID-19 virus, impacted positively in controlling the transmission of infective conjunctivitis as well.

However, the reduction in cases of infective conjunctivitis could be confounded by the fact that many patients might have refrained from visiting the hospital during the pandemic either due to fear of contracting COVID-19 or due to mobility restrictions imposed during those times. As a control, we looked at the number of cases of corneal foreign bodies presenting to our hospital during the corresponding period, which contrarily had increased. Werkl et al. also reported a similar result (9). The surge in corneal foreign body cases at our center during the pandemic could be attributed to the closure of the majority of other clinics during that period. Nevertheless, this implies that the eye care-seeking behavior of the patients did not change much despite the challenges of the pandemic period.

Similar to our study, in a single academic center in the US, the cases of non-allergic conjunctivitis presenting to emergency department was noted to fall by 37.3% and in the same study the internet search interest in US for conjunctivitis also decreased by 34.2% after widespread implementation of public health interventions to mitigate COVID-19 (2). Similarly in a Spanish primary level hospital, the ophthalmological emergencies reduced by 37.4% (13).

Our data also show significant changes in the annual distribution pattern of infective conjunctivitis during the pandemic period. Prior to and after the COVID-19 era, the prevalence of infective conjunctivitis was higher during the months of June through October. However, this pattern was not observed during the pandemic. Similar trend was observed in a study wherein the highest number of cases of adenoviral conjunctivitis was noted from July to September (14).

Our results also have shown that infective conjunctivitis affects the adult males more. Similar preponderance in adult male gender was observed in a study by Shrestha SP. et al (15). This result indicates that infective conjunctivitis is more prevalent among mobile and productive working age groups, which may translate into the economic burden associated with this condition.

The results of our study might suggest that the preventive measures implemented to restrained COVID-19 disease can also be applied to prevent the spread of infective conjunctivitis. And the data about the annual distribution pattern of infective conjunctivitis can help policy makers to implement effective countermeasures.

In conclusion, according to our data, the total number of cases of infective conjunctivitis decreased significantly during the COVID-19 pandemic along with the change in the annual distribution pattern. This reduction in the number and the change in the pattern of presentation may be attributed to the widespread implementation of preventive public health measures during the pandemic period.

## Data Availability

All relevant data are within the manuscript and its Supporting Information files.

## Acknowledgements

We would like to extend our gratitude to Mr. Sudhatma Karki and Mr. Jenish Gorkhali from the Information Technology (IT) Department of Tilganga Institute of Ophthalmology for facilitating us with the necessary data of the patients for this study.

## Financial disclosure

There is no financial disclosure to me made with regard to this study.

## Notes

### Competing Interest Statement

The authors have declared no competing interest.

### Funding Statement

The author(s) received no specific funding for this work.

### Author Declarations

This study was approved by the Institutional Review Committee of Tilganga Institute of Ophthalmology with approval no. Ref:20/2022 dated 13/11/2022.

## References

1. Alfonso SA, Fawley JD, Lu XA. Conjunctivitis. Prim Care - Clin Off Pract. 2015;42(3):325–45.

2. Lavista Ferres JM, Meirick T, Lomazow W, Lee CS, Lee AY, Lee MD. Association of Public Health Measures during the COVID-19 Pandemic with the Incidence of Infectious Conjunctivitis. JAMA Ophthalmol. 2021;98104:1–7.

3. Ramirez DA, Porco TC, Lietman TM, Keenan JD. Epidemiology of conjunctivitis in US emergency departments. JAMA Ophthalmol. 2017;135(10):1119–21.

4. Nauheim RC, Romanowski EG, Araullo-Cruz T, Kowalski RP, Turgeon PW, Stopak SS, et al. Prolonged Recoverability of Desiccated Adenovirus Type 19 from Various Surfaces. Ophthalmology [Internet]. 1990;97(11):1450–3. Available from: 10.1016/S0161-6420(90)32389-8

5. Azar MJ, Dhaliwal DK, Bower KS, Kowalski RP, Gordon YJ. Possible consequences of shaking hands with your patients with epidemic keratoconjunctivitis. Am J Ophthalmol [Internet]. 1996;121(6):711–2. Available from: 10.1016/S0002-9394(14)70640-3

6. Shekhawat NS, Shtein RM, Blachley TS, Stein JD. Antibiotic Prescription Fills for Acute Conjunctivitis among Enrollees in a Large United States Managed Care Network. Ophthalmology [Internet]. 2017;124(8):1099–107. Available from: 10.1016/j.ophtha.2017.04.034

7. Lee CS, Lee AY, Akileswaran L, Stroman D, Najafi-Tagol K, Kleiboeker S, et al. Determinants of Outcomes of Adenoviral Keratoconjunctivitis. Ophthalmology. 2018;125(9):1344–53.

8. Olsen SJ, Azziz-Baumgartner E, Budd AP, Brammer L, Sullivan S, Pineda RF, et al. Decreased influenza activity during the COVID-19 pandemic—United States, Australia, Chile, and South Africa, 2020. Am J Transplant [Internet]. 2020;20(12):3681–5. Available from: 10.1111/ajt.16381

9. Werkl P, Hoeflechner L, List W, Lindner E. The impact of social distancing on conjunctivitis cases—a retrospective single-center observation report. Graefe’s Arch Clin Exp Ophthalmol [Internet]. 2021;(0123456789):10–2. Available from: 10.1007/s00417-021-05392-w

10. Güemes-villahoz N, Burgos-blasco B, García-feijoó J, Sáenz-francés F, Arriola-villalobos P, Martinez-de-la-casa JM, et al. Conjunctivitis in COVID-19 patients : frequency and clinical presentation. 2020;2501–7.

11. Sen M, Honavar SG, Sharma N, Sachdev MS. COVID-19 and Eye: A Review of Ophthalmic Manifestations of COVID-19 PMID: Quick Response Code. 2021 [cited 2022 Aug 20]; Available from: https://www.ijo.in

12. Wu P, Luo C, Liu Q, Qu; Xingguang, Liang L, Wu K. Characteristics of Ocular Findings of Patients With Coronavirus Disease 2019 (COVID-19) in Hubei Province, China. JAMA Ophthalmol [Internet]. 2020;138(5):575–8. Available from: https://jamanetwork.com/

13. González-Martín-Moro J, Guzmán-Almagro E, Izquierdo Rodríguez C, Fernández Hortelano A, Lozano Escobar I, Gómez Sanz F, et al. Impact of the COVID-19 Lockdown on Ophthalmological Assistance in the Emergency Department at a Spanish Primary Level Hospital. J Ophthalmol. 2021;2021:1–9.

14. Lee J, Bilonick RA, Romanowski EG, Kowalski RP. Seasonal Variation in Human Adenovirus Conjunctivitis: A 30-Year Observational Study. Ophthalmic Epidemiol [Internet]. 2018;25(5–6):451–6. Available from: 10.1080/09286586.2018.1509096

15. Shrestha SP, Khadka J, Pokhrel AK, Sathian B. Acute bacterial conjunctivitis - antibiotic susceptibility and resistance to commercially available topical antibiotics in Nepal. Nepal J Ophthalmol. 2016;8(15):23–35.

